# Genomic discovery and functional validation of MRP1 as a novel fetal hemoglobin modulator and potential therapeutic target in sickle cell disease

**DOI:** 10.1101/2023.03.14.23287244

**Authors:** Yannis Hara, Emily Kawabata, Viktor T. Lemgart, Paola G. Bronson, Alexandra Hicks, Robert Peters, Sriram Krishnamoorthy, Jean-Antoine Ribeil, Lisa J. Schmunk, Jennifer Eglinton, Nicholas A. Watkins, David J. Roberts, Emanuele Di Angelantonio, John Danesh, William J. Astle, Dirk S. Paul, Samuel Lessard, Adam S. Butterworth

**Author notes:** These authors contributed equally to this work (Yannis Hara, Emily Kawabata, William J. Astle, Dirk S. Paul, Samuel Lessard, Adam S. Butterworth). Correspondence to Professor Adam S. Butterworth.

## Abstract

Sickle cell disease (SCD) remains a major health burden with limited treatment options. Despite promising gene-editing clinical trials, there is an unmet need for cost-effective therapies. As induction of fetal hemoglobin (HbF) is an established therapeutic strategy for SCD, we conducted a genome-wide association study of circulating HbF levels in ~11,000 participants to identify further HbF modulators. We identified associations in 11 genomic regions, including eight novel loci such as *ABCC1* (encoding multidrug resistance-associated protein 1, MRP1). Using gene-editing and pharmacological approaches, we showed that inhibition of MRP1 increases HbF, intracellular glutathione levels, and reduces sickling in erythroid cells from SCD patients. Overall, our findings identify several novel genetically-validated potential therapeutic targets for SCD, including promising proof-of-principle results from small molecule inhibition of MRP1.

## INTRODUCTION

Sickle cell disease (SCD) is a common monogenic disease and major health burden. There are an estimated 20 million patients worldwide, predominantly in sub-Saharan Africa, where up to 50% of those affected die in childhood (Ranque et al, 2022). SCD has high healthcare costs (>$1 billion per year for <100,000 patients in the US alone) and effective treatment options are limited (Johnson et al, 2022). SCD is caused by a single amino acid substitution in *HBB*, encoding adult hemoglobin (HbA) in red blood cells (RBCs). Homozygosity for this substitution leads to polymerization of the hemoglobin tetramer and deformation of RBCs. This ‘sickling’ of RBCs leads to hemolytic anemia, vascular occlusion associated with painful crises, organ dysfunction, and shortened lifespan (Telen et al, 2019; Steinberg et al, 2022).

Fetal hemoglobin (HbF), in which the hemoglobin β-chain is encoded by the γ-globin genes (*HBG1*, *HBG2*), is highly expressed at the fetal developmental stage and is predominantly replaced by HbA following birth through a process known as hemoglobin switching. However, residual levels of HbF (typically <1%) remain in adults, particularly in SCD patients in whom HbF levels may reach 10%. Higher levels of HbF in SCD patients are associated with lower mortality and morbidity because HbF inhibits the polymerization of sickle hemoglobin by excluding the mutant *HBB* protein from the hemoglobin polymer, which leads to increased RBC lifespan and efficiency of erythropoiesis (Platt et al, 1994; El Hoss et al, 2021). Indeed, SCD patients with substantially elevated HbF levels, such as those who also have the rare monogenic condition hereditary persistence of fetal hemoglobin (HPFH), can appear clinically asymptomatic. Inducing HbF production is, therefore, an established therapeutic strategy for SCD, as well as other β-hemoglobinopathies.

Hydroxyurea, an anti-metabolite which raises HbF levels, is the most widely used treatment option for SCD. However, there is still unmet need for alternate therapies as hydroxyurea is only effective in around 70% of patients and carries a boxed warning for immune suppression and cancer (Lanzkron et al, 2008). The ability of gene therapy to correct autologous hematopoietic stem cells (HSCs) offers a potential cure for SCD. For example, early-phase clinical trials have used gene-editing to silence *BCL11A* in patient-derived HSCs, resulting in up-regulation of HbF and reduced clinical manifestations of SCD (Esrick et al, 2021; Frangoul et al, 2021; Alavi et al, 2022). However, even if successful in late-stage clinical trials, the high cost of gene therapy would likely prevent access in areas with the highest SCD prevalence, including Africa and India (Hoban et al, 2016; Bourzac, 2017).

The discovery of BCL11A as a promising target for SCD arose from genome-wide association studies (GWAS) of circulating HbF levels, which identified the *BCL11A*, *HBB*, *HBS1L-MYB*, and *NFIX* loci (Uda et al, 2008; Lettre et al, 2008; Danjou et al, 2015). The associated alleles at *BCL11A* disrupt an enhancer site that prevents *BCL11A* expression in the erythroid lineage when deleted (Sankaran et al, 2008; Bauer et al, 2013, Canver et al, 2015). *BCL11A* encodes B-cell lymphoma/leukemia 11A, a zinc-finger protein that acts as a transcriptional repressor of the γ-globin genes. Therefore, disrupting *BCL11A* (or its enhancers) is a compelling therapeutic strategy for up-regulating HbF in SCD, as well as other β-hemoglobin disorders such as β-thalassemia.

The success in translating HbF-associated variants identified by GWAS into understanding of disease mechanisms and novel therapeutic strategies suggests that additional targets could be identified for SCD following a similar approach. Targets that could be modulated through small molecules may be particularly attractive given the current challenges for gene therapies. Here, we conduct a GWAS of circulating HbF levels in ~11,000 participants. We identify several new associations, including a low-frequency missense variant in *ABCC1*, which encodes multidrug resistance-associated protein 1 (MRP1). Using gene-editing and pharmacological approaches, we identify MRP1 as a novel HbF regulator and anti-sickling mechanism acting through NRF2-mediated oxidative stress signaling, establishing MRP1 as a potential novel therapeutic target in SCD.

## RESULTS

### Genome-wide association study of HbF levels in the INTERVAL study

We conducted a GWAS of peripheral blood HbF levels in 11,004 generally healthy blood donors participating in the INTERVAL study [<u>Supplementary Table 1</u>] (Di Angelantonio et al, 2017). The estimated genomic control inflation factor was 1.047, suggesting little genome-wide evidence for confounding [<u>Supplementary Figure 1</u>]. Single-variant analysis identified ten genomic regions containing alleles associated with HbF levels at genome-wide significance (*P*<5×10^−8^) [<u>Supplementary Figure 2</u>; <u>Supplementary Table 2</u>], including the previously identified loci near the *BCL11A*, *HBS1L-MYB*, and *HBB* genes (Uda et al, 2008; Lettre et al, 2008; Danjou et al, 2015). However, we did not have good power to test the associations with previously reported variants in the *NFIX* locus as they were much rarer (minor allele frequency [MAF]~0.001) in the UK-based INTERVAL study than the original Sardinian discovery population (MAF~0.03) (Danjou et al, 2015). We identified seven loci not previously associated with HbF levels near *ABCC1*, *BACH2*, *GCDH, GPR75-ASB3*, *GRIK2*, *PFAS*, and *TMEM161B*. Stepwise genome-wide conditional analysis identified eight further conditionally independent associations, including an additional novel locus (*ARHGAP39*) and seven secondary signals at *ABCC1*, *BCL11A*, *HBB*, *HBS1L-MYB,* and *PFAS* [Table 1].

**Table 1.**
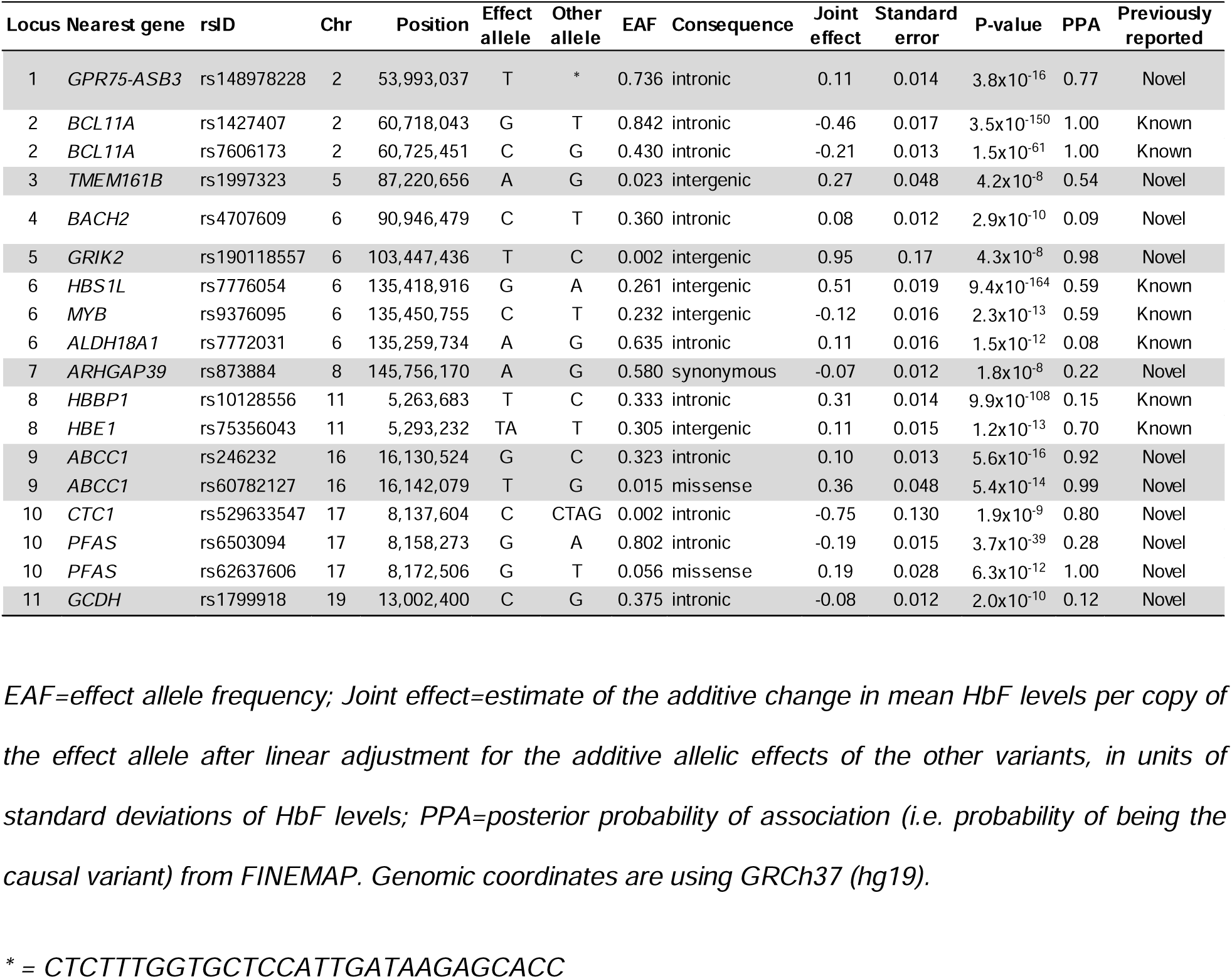
18 conditionally independent associations with HbF at eleven genomic loci.

Sentinel variants were predominantly intronic or intergenic, with the exceptions of a common synonymous variant in *ARGHAP39* (rs873884; MAF=0.42), and low-frequency missense variants in *PFAS* (rs62637606, p.Phe1314Cys; MAF=0.06) and *ABCC1* (rs60782127, p.Arg433Ser; MAF=0.02). Our sample size also allowed the detection of associations with two rare variants, an intronic variant in *CTC1* at the *PFAS* locus (rs529633547; MAF=0.002) and an intergenic variant in a gene desert on chromosome 6 (rs190118557; MAF=0.002).

As several of these HbF-associated variants are known to be associated with total hemoglobin concentration and other red cell parameters from previous GWAS meta-analyses [<u>Supplementary Table 3</u>] (Chen et al, 2020), we used the detailed complete blood count data available in the INTERVAL study to test for mediation. The data included 30 RBC traits that reflect the counts or biochemical and structural properties of RBCs at various stages of their lifespan [<u>Supplementary Table 4</u>]. Adjustment for these RBC traits did not attenuate the effects of the variants on HbF <u>[Supplementary Table 5</u>], suggesting the effects of these variants on HbF levels are not driven by changes in these RBC parameters.

For nine of the 18 associations, we were able to identify variants with a high probability of causality (posterior probability [PP]>0.70) [Table 1]. These included several previously reported signals, such as the two functional *BCL11A* enhancer variants (rs1427407 [PP=1.0] and rs7606173 [PP=1.0]) (Bauer et al, 2013) and rs75356043 (PP=0.70) near the hemoglobin gene cluster on chromosome 11. We also identified highly probable causal variants at novel loci, such as rs62637606 (PP=1.0) in *PFAS*, rs529633547 (PP=0.80) in *CTC1* and both the low-frequency missense variant p.Arg433Ser (rs60782127 [PP=0.99]) and a common intronic variant (rs246232 [PP=0.92]) in *ABCC1* (pairwise *r^2^*~0.003, *D’*=1 in European ancestries).

### Molecular phenome-scan of *ABCC1*-p.Arg433Ser confirms the role of MRP1 in glutathione metabolism

We chose to prioritize an association at the *ABCC1* locus for elucidation of the causal mechanism because the strong evidence from statistical fine-mapping implicating the low-frequency, missense variant in *ABCC1* suggested the likely causal variant and gene for this association.

*ABCC1* encodes MRP1, one of nine drug-transporting ATP-binding cassette transporters. To assess the broad phenotypic consequences of genetic perturbation of *ABCC1*, we conducted a molecular phenome scan using metabolomic and proteomic data derived from blood samples of INTERVAL participants. The strongest plasma metabolite associations for *ABCC1*-433Ser (rs60782127-T) were with lower levels of metabolites from the glutathione metabolism pathway, consistent with reduced cellular efflux of glutathione metabolites, as well as the gamma-glutamyl amino acid and acylcarnitine pathways [<u>Supplementary Table 6</u>, <u>Supplementary Figure 3</u>]. Of ~3500 plasma proteins tested, the protein with the strongest evidence for association was glutathione S-transferase omega 1 (GSTO1) (additive effect per copy of the 433Ser allele=0.50 SD, *P*=1.9×10^−6^) [<u>Supplementary Figure 3</u>]. A broader phenome scan of quantitative traits and disease phenotypes using several biobanks and published GWAS showed associations with only blood cell traits and anthropometric measures, which were all weak in magnitude [<u>Supplementary Figure 4</u>; <u>Supplementary Table 7</u>]. These findings support p.Arg433Ser as the likely causal variant (and *ABCC1* as the likely causal gene) at this locus, and support the role of MRP1 in glutathione metabolism *in vivo*.

### *ABCC1*-433Ser leads to increased HbF levels in HUDEP-2 cells

To evaluate the potential role of *ABCC1*-433Ser in HbF expression, we generated erythroid cell line (HUDEP-2) clones that were homozygous for the *ABCC1*-433Ser allele (N=3) using CRISPR-Cas9 (Kurita et al, 2013). We observed increased intracellular total glutathione levels (16-fold increase in mean, *P*<0.01) in clones carrying 433Ser compared to wild-type cells [Figure 1a]. These clones also produced significantly more HbF-positive cells (F-cells; 5.7-fold increase in mean across replicates, *P*<0.01) and HbF as shown by flow cytometry analysis, HPLC (8.8% of total globin in clone C6 vs 0.2% in wild-type cells), and western blot [Figures 1b,c,d]. The effect of 433Ser on HbF induction was further confirmed in additional clones D2 and E5 [Figure 1b, <u>Supplementary Figure 5</u>]. These results validate the impact of *ABCC1*-p.Arg433Ser on glutathione efflux and HbF induction.

**Figure 1.**
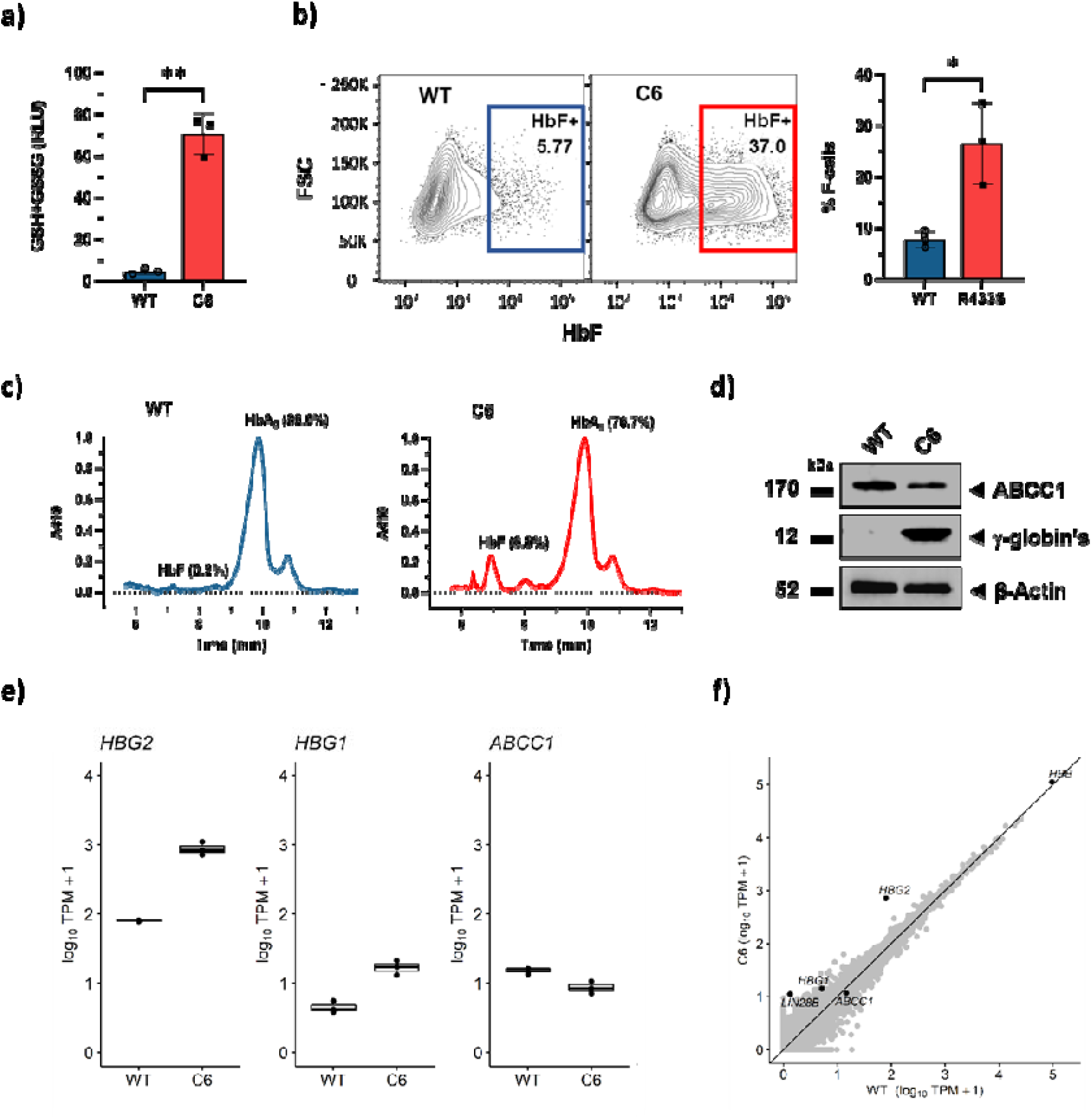
*ABCC1-*433Ser knock-in leads to increased HbF in HUDEP-2 cells. a) Increase in total intracellular glutathione (GSH+GSSG) levels in HUDEP-2 clone C6 homozygous for the 433Ser allele (N=3) compared to wild-type cells (N=3) confirms ABCC1 loss-of-function. Two-tailed t-test, **P<0.01. RLU: relative light units. b) Homozygous knock-in of the ABCC1-433Ser allele in HUDEP-2 clone C6 (N=3) leads to increased HbF levels compared to wild-type cells (N=3) as quantified by flow cytometry after 7 days of erythroid differentiation (<u>right</u>). Two-tailed t-test, *P<0.05. Representative flow cytometry plots from a single experiment showing increased HbF in HUDEP-2 C6 clones (<u>left</u>). FSC: Forward scatter. c) HbF induction in HUDEP-2 clone C6 homozygous for the 433Ser allele was confirmed by cation-exchange high-performance liquid chromatography (HPLC). Representative example from a single experiment. Similar results were obtained for three independent experiments. 410 = Absorbance 410 nm. d) Western blot shows increased gamma-globin in HUDEP-2 clone C6 compared to wild-type cells, as well as a reduction in MRP1 protein levels. Representative example from a single experiment. β-actin served as an internal control. e) Transcriptomic analysis of 433Ser HUDEP-2 cells confirms up-regulation of gamma-globin genes and down-regulation of ABCC1. Differential gene expression analysis between ABCC1-433Ser homozygous clone C6 (N=3) and wild-type HUDEP-2 cells (N=3). Replicates are from three independent experiments. Figure shows normalized log_10_ counts of HBG2, HBG1, and ABCC1 in clone C6 and wild-type HUDEP-2 cells. f) HUDEP-2 433Ser induces transcriptional changes. Plot shows gene expression levels in wild-type HUDEP-2 cells averaged across replicates (N=3, x-axis) compared to genes in clone C6 (N=3, y-axis). Gene expression levels were measured using RNAseq and are reported as transcripts per million (TPM).

Using RNA-sequencing, transcriptomic analysis of clone C6 confirmed increased expression of the γ-globin genes compared to wild-type cells (*HBG2*, 12.0-fold increase, *P*_adj_=5×10^−43^; *HBG1*, 4.2-fold increase in mean, *P*_adj_=8×10^−10^) [Figure 1e, <u>Supplementary Table 8</u>]. We also observed decreased *ABCC1* expression (0.6-fold decrease in mean, *P*_adj_=3×10^−6^) [Figure 1e], which was detectable by western blot at the protein level [Figure 1d]. Interestingly, clone C6 had increased expression of *LIN28B*, a gene highly expressed in fetal erythroblasts and previously implicated in HbF regulation [Figure 1f] (Basak et al, 2020; Lee et al, 2013; Lessard et al, 2018). Finally, differentially expressed genes were significantly enriched in the glucocorticoid receptor signaling pathway (*P*=1.9×10^−6^) [<u>Supplementary Table 9</u>].

To confirm that MRP1 is causally implicated in HbF production, we performed CRISPR-Cas9 gene editing to create *ABCC1* knockouts in HUDEP-2 cells. HUDEP-2 cells transfected with single guide RNAs (sgRNAs) against *ABCC1* showed a strong increase in total intracellular glutathione levels (the sum of reduced glutathione [GSH] and oxidized glutathione [GSSG]) (6-fold increase in mean across replicates, *P*<0.05) [<u>Supplementary Figure 6a</u>], consistent with reduced efflux of these metabolites. We also saw a significantly higher number of F-cells (2.5-fold increase in mean across replicates, *P*<0.05), as well as increased γ-globin production (5.6% of total globin in *ABCC1* knockout cells vs 0.1% in non-targeting sgRNA) after erythroid differentiation compared to control, confirming the role of MRP1 in HbF induction [<u>Supplementary Figure 6b</u>]. This increase in HbF level was ascertained by both high-performance liquid chromatography (HPLC) and western blot [<u>Supplementary Figures 5c,d</u>].

We next sought to confirm that disruption of *ABCC1* leads to increased HbF production in human CD34+ erythroid cells. First, we used CRISPR-Cas9 to abrogate *ABCC1* in CD34+ cells. Similar to HUDEP-2 cells, sgRNAs targeting *ABCC1* led to an increase in total intracellular glutathione (8-fold increase in mean across replicates, *P*<0.01) [<u>Supplementary Figure 7a</u>], F-cells (1.8-fold increase in mean across replicates, *P*<0.05) and HbF production (5.9% of total globin in *ABCC1* knockout cells vs 0.1% in non-targeting sgRNA) after 14-day erythroid differentiation [<u>Supplementary Figures 7b,c,d</u>].

### Pharmacological inhibition of MRP1 increases HbF expression and prevents sickling in differentiated CD34+ cells from SCD patients

MK571 is a known selective leukotriene D4 receptor antagonist and inhibitor of MRP1 that was developed as a potential bronchodilator for asthma patients but subsequently discontinued (Gekeler et al, 1995). Differentiation of CD34+ HSCs in the presence of MK571 caused an increase in total glutathione (6.5-fold increase, *P*<0.01) [Figure 2a], in F-cells (1.8-fold increase, *P*<0.05) and HbF production (8.5% of total globin in MK571-treated cells vs 0.2% in DMSO-treated cells) [Figure 2b,c,d]. Importantly, we observed a similar increase in F-cells after MK571 treatment of CD34+ HSCs from SCD patients (1.9-fold increase, *P*<0.05) [Figure 2e]. Furthermore, these cells resist sickling triggered by hypoxia (2% O_2_) compared to controls [Figure 2f,g], suggesting that inhibition of MRP1 could prevent sickling in SCD patients.

**Figure 2.**
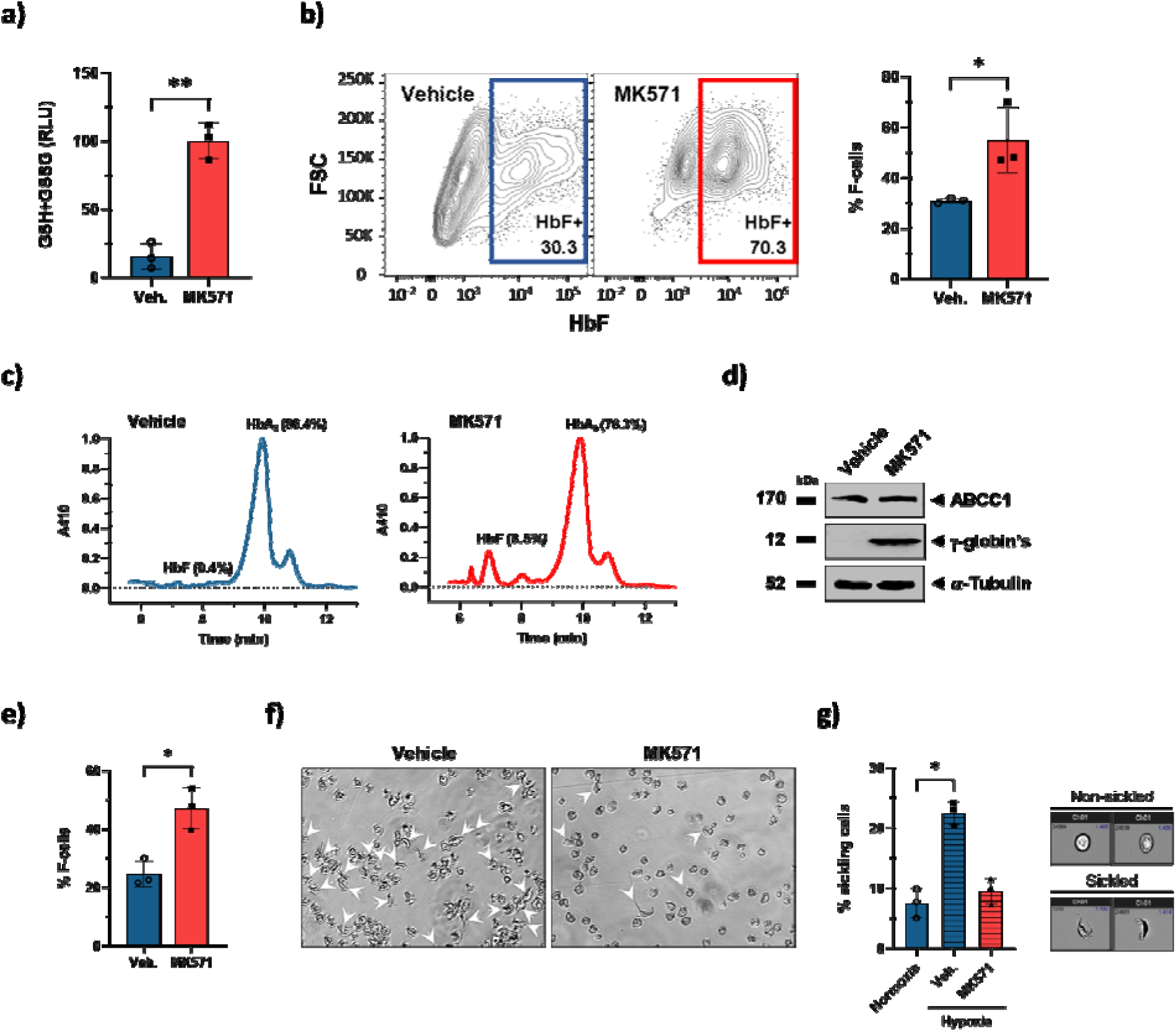
Pharmacological inhibition of MRP1 in erythroid cells derived from CD34+ HSCs. a) Increase in total intracellular glutathione levels in MK571-treated CD34+ erythroid cells (N=3) compared to vehicle (0.1% DMSO) (N=3) confirms ABCC1 target engagement by MK571. Two-tailed t-test, **P<0.01. RLU: relative light units. b) MRP1 inhibition by MK571 (50µM) significantly increases HbFIZlpositive cells by day 14 in differentiating CD34+ cells from healthy donors compared to vehicle (0.1% DMSO) as quantified by flow cytometry (<u>right</u>). *P<0.05. Panels are representative plots from a single donor (<u>left</u>). FSC: Forward scatter. c) HbF induction in MK571-treated CD34+ cells was confirmed by cation-exchange high-performance liquid chromatography (HPLC). 410 = Absorbance 410 nm. d) Representative western blot of differentiating CD34+ cells at day 14 confirms HbF induction in MK571-treated CD34+ cells compared to vehicle (0.1% DMSO). Similar results were obtained for three independent donors. α-tubulin served as an internal control. e) MRP1 inhibition by MK571 (50µM) significantly increases HbFIZlpositive cells by day 14 in differentiating CD34+ cells from sickle donors compared to vehicle (0.1% DMSO) as quantified by flow cytometry. *P<0.05. f) Representative microscopic images of 21-day differentiated CD34+ cells from sickle donors under hypoxia (4 hours) after treatment with MK571 (50µM) or vehicle (0.1% DMSO) confirms resistance to sickling due to MRP1 inhibition (all magnifications 20x). Arrows indicate abnormally shaped cells. Similar results were obtained for three independent donors. g) HbF induction in MK571-treated differentiated CD34+ erythroid cells from sickle donors protects cells from sickling under hypoxia. CD34+ cells from SCD donors were differentiated for 21 days in the presence of MK571 (50µM, N=3) or DMSO (0.1%, N=3) and then exposed to hypoxia for 4 hours to induce sickling. Quantification of normal and sickled cells under normoxia or hypoxia was quantified by Sickle cell Imaging Flow Cytometry Assays (SIFCA) (<u>left</u>). Two-tailed t-test, *P<0.05 (N=3). Representative images by SIFCA of the cell populations under normoxia gated as non-sickled, or under hypoxia (2% oxygen) gated as sickled (all magnifications 60×) (<u>right</u>). Mean ± SEM (N=3 independent HbSS SCD donors) and *P<0.05 (two-tailed t-test).

To rule out impaired erythroid differentiation as a mechanism explaining the observed effects of MRP1 inhibition on HbF induction, we measured enucleation of terminally differentiated MK571-treated CD34+ HSCs by flow cytometry at day 21 of differentiation. We did not observe any significant difference between MK571 and DMSO-treated cells [<u>Supplementary Figure 8a</u>]. Consistently, we did not detect a significant effect of MK571 treatment on either the expression of CD235a, measured by flow cytometry on day 21 of differentiation [<u>Supplementary Figure 8b</u>], or on the expression of other erythroid differentiation markers, including GATA1, BAND-3 (SLC4A1), LRF, and ALAS2, measured by western blot on day 15 of differentiation [<u>Supplementary Figure 8c</u>]. Finally, Giemsa staining of CD34+ cells did not reveal differences in cell morphology between MK571-treated CD34+ cells and control [<u>Supplementary Figure 8d</u>]. Taken together, these data confirm the role of MRP1 in HbF induction without affecting erythroid differentiation.

### Inhibition of MRP1 activates NRF2-activated oxidative stress response

To identify the mechanism by which MRP1 modulates HbF, we measured the transcriptome, proteome, and phospho-proteome of CD34+ cells exposed to DMSO or MK571. We observed increased gene expression of *HBE1*, encoding ε-(embryonic-) globin (13.8-fold increase, *P*=2×10^−17^, *P*_adj_=1×10^−13^) [<u>Supplementary Table 10</u>]. Further, we detected an increase in γ-globin and ε-globin production at the protein level, after normalizing for the sum of the expression of β-like globins to account for the variability in total globin expression levels (*P*<0.05) [Figure 3a]. The gene and protein expression levels were broadly in agreement, with a positive correlation in fold-changes between DMSO- and MK571-treated cells (*r*=0.3, *P*=6×10^−151^) [Figure 3b]. We observed a strong up-regulation of RRAD (encoding the Ras Related Glycolysis Inhibitor and Calcium Channel Regulator) in both the transcriptomic (39.9-fold increase, *P*_adj_=9×10^−23^) and proteomic analysis (11.4-fold increase, *P*_adj_=0.02). No protein displayed differential phosphorylation after correction for multiple testing [Figure 3c].

**Figure 3.**
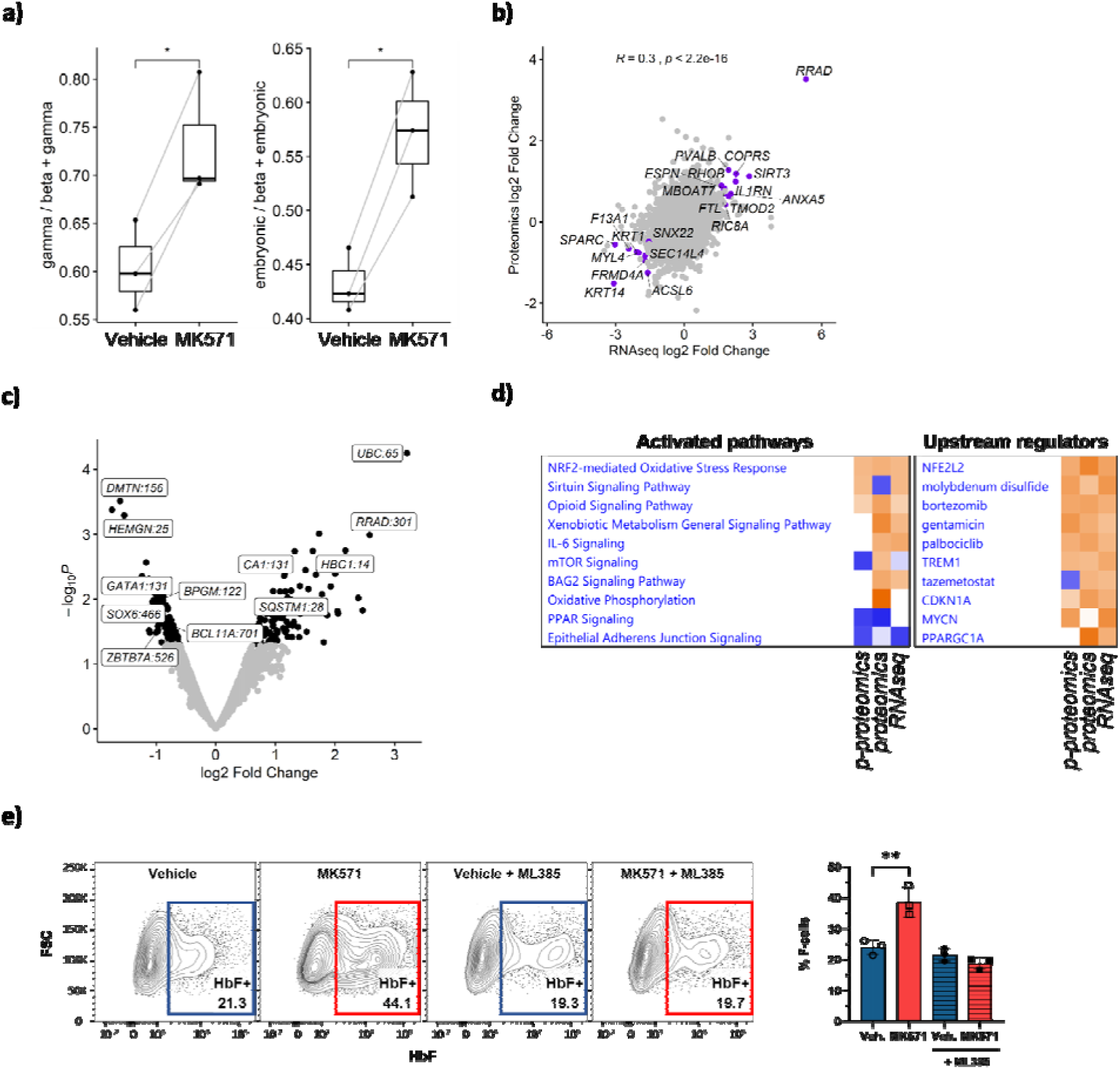
The NRF2 pathway is enriched in erythroid cells derived from MK571-treated CD34+ HSCs. a) Increased expression of gamma- and embryonic-globins in MK571-treated CD34+ HSCs compared to control (0.1% DMSO). Protein expression was quantified by mass spectroscopy via tandem mass tag (TMT) labeling. Protein levels are normalized on the sum of beta-like globins. N=3 independent donors; *P<0.05. b) Correlation between differential protein and gene expression log_2_ fold changes as measured by TMT proteomics and RNAseq. Purple points show genes with differential gene expression false discovery rate Q-value < 0.05 and proteins with protein differential expression Q-value < 0.25. Differential gene expression analysis was performed using DESeq2. Differential protein expression analysis was performed using DEqMS. Correlation was assessed using Pearson’s correlation. c) Volcano plot showing differentially phosphorylated proteins measured by TMT mass spectroscopy. Black points show protein sites with a differential phosphorylation nominal P<0.05. d) Top 10 pathways and upstream regulators consistently enriched in the RNAseq, proteomics, and phospho-proteomics analysis. Heatmap shows pathway enrichment Z-scores with positive values in red and negative values in blue. P-proteomics: phospho-proteomics. e) HbF induction by MK571 is abolished in the presence of NRF2 inhibitor ML385 (10µM), suggesting a role of NRF2 in HbF induction triggered by ABCC1 inhibition (<u>right</u>). Panels are representative plots from a single donor (<u>left</u>). Two-tailed t-test, *P<0.05, N=3.

We performed a comparative analysis of enriched pathways and upstream regulators from the transcriptomic, proteomic, and phospho-proteomic experiments using Ingenuity Pathway Analysis (IPA). This analysis revealed a consistent induction of the NRF2-mediated oxidative stress response pathway [Figure 3d]. We found NRF2 (encoded by *NFE2L2*) to be enriched as an upstream regulator in all pathway analyses. To confirm the role of NRF2 in MRP1 inhibition-mediated HbF up-regulation, we treated CD34+ HSCs with MK571 in the presence of ML385, a selective NRF2 inhibitor (Singh et al, 2016). In the presence of ML385, MK571-induced HbF up-regulation was abolished [Figure 3e], while cell differentiation was not affected (data not shown), suggesting that NRF2 mediates HbF induction when MRP1 is inhibited.

## DISCUSSION

Prior discovery of associations in *BCL11A* with HbF levels and subsequent functional follow-up to elucidate the causal mechanism represent a compelling example of translating genetic discoveries into therapeutic targets, with promising initial results in clinical trials (Esrick et al, 2021; Frangoul et al, 2021; Alavi et al, 2022). To identify additional therapeutic targets for SCD using this approach, we conducted a GWAS of peripheral blood HbF levels, involving over 11,000 participants. We extend the number of HbF-associated regions from three established loci (*BCL11A*, *HBB*, *HBS1L-MYB*) to eleven. Some of the novel loci include genes with established biological links to HbF, such as *KLF1* (Kruppel-like factor 1), an erythroid transcription factor that binds to and activates the promoter of *BCL11A* (Zhou et al, 2010). The remaining loci represent starting points for understanding novel regulators of HbF that could lead to the identification of further, much needed, therapeutic targets for SCD.

Due to the presence of a low-frequency missense variant in *ABCC1* (p.Arg433Ser), we selected this locus for functional follow-up and pharmacological perturbation to elucidate a new regulatory pathway for HbF. The 433Ser allele, which is predicted to be deleterious by computational tools such as SIFT and PolyPhen-2, affects a conserved residue located within a cytoplasmic loop of a membrane-spanning domain of MRP1 (Karczewski et al, 2020). Although present at low-frequency in European ancestry populations, the allele is rare (MAF 0.001-0.004) in African and South Asian ancestries, and absent in East Asian populations. Using CRISPR-Cas9 and gene-edited clones, we observed effects of the 433Ser allele and abrogation of the *ABCC1* gene on the regulation of HbF at the gene and protein level in both an erythroid cell line and primary erythroid differentiated CD34+ HSCs, validating both the causal variant and effector gene/protein at the locus. Small molecule inhibitors of MRP1 have been developed including MK571, a selective leukotriene D4 receptor antagonist, and Reversan, a cell-permeable pyrazolopyrimidine. Treatment of CD34+ cells with MK571 increased HbF levels, confirming that small molecule inhibition of MRP1 can reactivate HbF. Although MK571 also inhibits Multidrug resistance-associated protein 4 (MRP4), our gene-editing experiments suggest that the HbF induction seen with MK571 treatment is due to MRP1 inhibition. MK571 treatment did not affect erythroid differentiation, suggesting that the effect of MRP1 on HbF modulation is not due to changes in the differentiation and maturation of erythroid cells.

The 433Ser allele in MRP1 is known to confer a 2-fold reduction in the transport of the GSH-conjugated metabolite leukotriene C4 and oestrone sulfate, with half the V_max_-value of wild-type MRP1 for these substrates (Conrad et al, 2002). Our molecular phenome scan showed associations of this allele with plasma GSTO1 protein levels and lower plasma levels of metabolites related to glutathione metabolism, confirming that the variant impacts glutathione-related metabolism *in vivo* and that 433Ser confers a reduction in MRP1 activity. Both CRISPR-Cas9-mediated knockdown of *ABCC1* in erythroid cells and clones homozygous for the 433Ser allele showed significant increases in total intracellular glutathione levels. We also found increases in total glutathione levels with CRISPR-Cas9-mediated knockdown of *ABCC1* and with MK571 treatment in erythroid differentiated CD34+ HSCs. This is consistent with previous studies showing that MRP1 inhibition maintains intracellular glutathione in other cell types and the role of MRP1 as an intracellular glutathione efflux pump. L-glutamine has been approved by the FDA for the treatment of sickle cell diseases based on a reduced frequency of vaso-occlusive crises in a phase 3 clinical trial (Niihara et al, 2018). L-glutamine is a non-essential amino acid that plays an important role in the pathophysiology of SCD as it contributes to the protection of erythrocytes from oxidative stress (Jafri et al, 2022; Sadaf & Quinn, 2020). Indeed, glutamine is a precursor of nicotinamide adenine dinucleotide (NAD+), a potent antioxidant in RBCs, as well as glutamate and subsequently glutathione. Therefore, it is hypothesized that glutamine supplementation in SCD can help replenish NAD+ and glutathione levels in RBC, protecting against oxidative stress and the resulting hemolysis (Jafrid et al, 2022; Morris et al, 2008; Morris et al, 2022; Sadaf & Quinn, 2020). However, this remains to be demonstrated clinically.

Importantly, we showed that pharmacological inhibition of MRP1 and subsequent HbF reactivation reduce sickling of SCD-derived erythroid cells under hypoxia. This confirms MRP1 inhibition as a potential therapeutic avenue to prevent complications of SCD, such as vaso-occlusive crises. MRP1 inhibition has previously been shown to reduce reactive oxygen species (ROS) and maintain intracellular glutathione levels, thereby protecting against oxidative stress (Nur et al, 2011a). SCD patients have a high burden of oxidative stress, in part due to hemolysis and chronic inflammation. In turn, oxidative stress in SCD can lead to endothelial dysfunction and acute inflammation. Higher levels of ROS have also been linked to increased rates of complications in SCD, including vaso-occlusive crises (Nur et al, 2011b). Transcriptomic analysis of the 433Ser homozygous clones revealed enrichment of genes involved in glucocorticoid receptor signaling, a pathway related to oxidative stress. MK571 treatment of RBCs from individuals with β-thalassemia reportedly reduces the formation of H_2_O_2_-induced free radicals due to increased intracellular glutathione (Muanprasat et al, 2013). This suggests that MRP1 inhibition reduces SCD complications by preventing sickling of RBCs and reducing oxidative stress, potentially via maintenance of intracellular glutathione.

Through transcriptomic, proteomic, and phospho-proteomics analyses, we demonstrated that MRP1 inhibition activates the NRF2 oxidative stress response pathway. Treatment of CD34+ cells with a specific NRF2 inhibitor, ML385, abolished MRP1-mediated induction of HbF, suggesting that MRP1 modulates HbF levels via NRF2. NRF2 activation increases HbF expression in multiple erythroid cell types, such as CD34+ cells, and reduces inflammation and anemia in transgenic SCD mouse models (Belcher et al, 2017; Ghosh et al, 2016; Krishnamoorthy et al, 2017). NRF2 positively modulates *HBG2* expression by binding to antioxidant response elements (ARE) elements located in its promoter (Macari et al, 2011, Zhu et al, 2017). Similarly, it has been reported that NRF2 binds ARE located in the *ABCC1* promoter to increase its transcription, resulting in an increase in MRP1 protein expression level (Ji et al, 2013; Chen et al, 2017; Hayashi et al, 2003). In addition, NRF2 activates genes involved in GSH synthesis and regeneration (Hanssen et al, 2021), tightly maintaining intracellular glutathione homeostasis. In our study, we showed that MRP1 inhibition activates NRF2, suggesting a feedback mechanism that counteracts the decrease in *ABCC1* expression (or MRP1 activity), with a subsequent increase in intracellular total glutathione levels [Figure 4].

**Figure 4.**
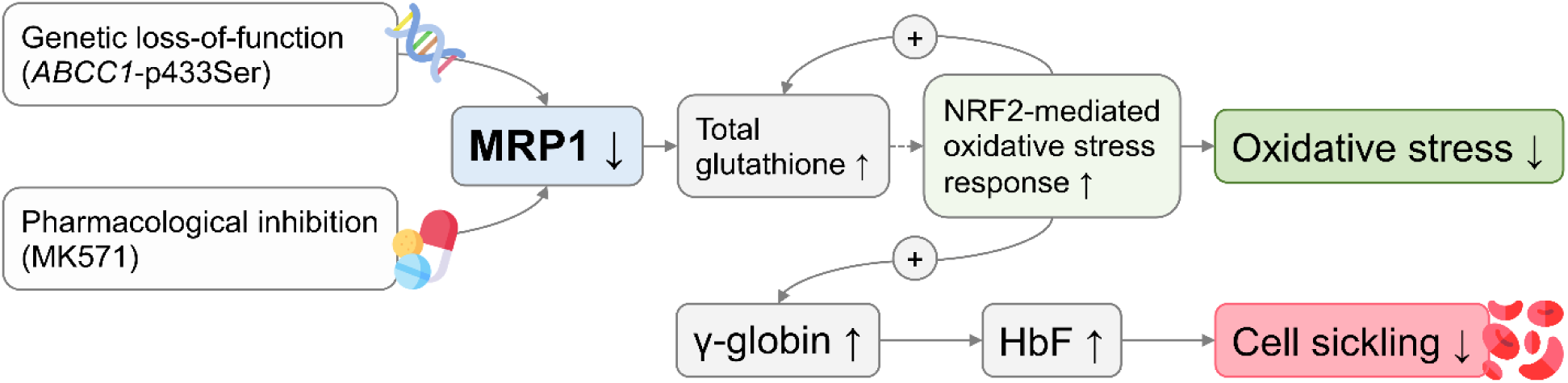
Proposed mechanism by which inhibition of MRP1 could be beneficial in SCD.

There is an unmet need for novel therapeutic targets that increase HbF in erythroid cells to develop safer and more accessible treatment options for SCD and other β-hemoglobin disorders (Steinberg et al, 2020). Our genome-wide discovery study and subsequent experimental follow-up suggest that MRP1 is a promising therapeutic target. Validation of MRP1 as a safe and efficacious treatment option for SCD will require *in vivo* testing to understand whether chronic inhibition of erythroid MRP1 to increase HbF expression significantly improves disease biomarkers, and to identify any potential safety concerns resulting from inhibition of the ubiquitous MRP1.

## METHODS

### Participants

We assayed HbF in 11,004 randomly selected participants from the INTERVAL BioResource (Moore et al, 2014; Di Angelantonio et al, 2017). The INTERVAL study involved ~50,000 generally healthy blood donors aged 18-80 recruited across England from 2012 to 2014. At recruitment, an EDTA tube of blood was collected before overnight transport to UK Biocentre (Stockport and Milton Keynes, UK) for centrifugation and aliquoting.

### HbF measurement

Whole blood samples from ~12,000 participants were shipped for assay between March 2017 and December 2017 to the University of Oxford. Plates were thawed overnight and processed on the day of assay. HbF was measured using mass spectrometry using the Bio-Rad VARIANT II automated assay system and was reported as a percentage of total hemoglobin, accurate to 1 decimal place.

The HbF level was adjusted for participant characteristics such as age, age^2^ and blood group, as well as technical factors (INTERVAL trial arm, the center at which the donor was recruited, the recruitment date, plate ID and assay run number). These factors were selected by stepwise regression to obtain a parsimonious set of technical variables explaining variance in the HbF levels. We also adjusted for the first 10 genetic principal components of ancestry to account for potential population stratification. In aggregate, these factors explained 32.6% of the variation in measured HbF levels. Residuals from the covariate adjustments were then normalized using a rank inverse normal transformation. After exclusions made because of duplicate samples, non-European ancestry, missing covariates or missing HbF levels, 11,004 participants remained for statistical analysis.

### Genotyping

DNA was extracted from whole blood aliquots at LGC Biosciences. Aliquots were shipped to ThermoFisher (formerly Affymetrix), Santa Clara, US for genotyping. We directly genotyped 820,967 variants using the Affymetrix UK Biobank array (Astle et al, 2016). After sample and variant QC, we imputed 87,696,888 variants using a combined 1000 Genomes/UK10K reference panel. Variants were filtered to remove those with low imputation quality (INFO<0.4) and MAF<0.002, leaving 14,910,742 variants for association testing.

### Genome-wide association analysis

We ran linear mixed models on the inverse normalized HbF levels for all autosomes using the BOLT-LMM v2.3 package (Loh et al, 2015; Loh et al, 2018). A conventional genome-wide significance threshold (*P*<5×10^−8^) was used.

To (a) identify secondary signals at the initial HbF-associated regions and (b) identify additional HbF-associated regions by removing variance from HbF levels explained by strongly associated genetic loci, we performed stepwise regional conditional analysis by conditioning on the lead variant in each region (+/- 1Mb from the lead variant). We initially conditioned on the lead variants from the three well-known loci (*BCL11A*, *HBS1L-MYB* and *HBB* loci). After six iterations, 18 conditionally independent signals at 11 loci reached genome-wide significance [Table 1]. To identify whether these associations were novel, we extracted the lead variants from previous GWAS of HbF (Uda et al, 2008; Lettre et al, 2008; Danjou et al, 2015) and considered associations as ‘novel’ if the r^2^ between our conditional lead variant and the previously reported lead variant was less than 0.04, a threshold based on visual inspection of regional association plots.

### Fine-mapping

Within each of the nine associated regions, we identified the most plausible set of causal variants using FINEMAP v1.3 (Benner et al, 2016). Under the assumption that the true causal variant has been measured in the locus, FINEMAP uses a shotgun stochastic search algorithm on the association summary statistics to calculate the posterior probability for the causality of each configuration of variants in a Bayesian framework. For each region (lead variant +/- 1Mb), we set the maximum number of causal variants to be the number of independent associations found from the conditional analysis. Other than the ‘corr_group’ parameter, which we set to 0.7, and the ‘n-iterations’ parameter, which we set to 10,000, we used the default values for all other parameters.

### Phenome-wide association analyses

To examine the associations of HbF-associated variants with molecular traits, phenotypes and diseases, we interrogated several publicly available resources (Open GWAS, GWAS Atlas, PhenoScanner [Staley et al, 2016], OpenTargets Genetics, eQTL Catalog) as well as specific databases including proteomic GWAS (ARIC, deCODE, Fenland, INTERVAL), metabolomic GWAS (INTERVAL) and gene expression GWAS (eQTLGen, GTEx).

### Reagents

MK571 and ML385 were purchased from Sigma Aldrich (cat#M7571 and #SML1833 respectively).

### CD34+ erythroid cell differentiation

We differentiated CD34+ cells in culture from the peripheral blood mononuclear cell (PBMC) fraction of healthy and SCD donor blood as described (Giarratana et al, 2005). Briefly, we cultured mobilized CD34+ human stem / progenitor cells (HSPC) from healthy individuals for 3 days in a maintenance media consisting of X-VIVO 10 (VWR cat#12002-004), 100 U/mL penicillin-streptomycin (ThermoFisher, cat#15140122), 2mM L-glutamine (Fisher Scientific, cat#25030-081), 100ng/mL Recombinant Human Stem Cell Factor (Thermofisher, cat#CTP2113), 100ng/mL Recombinant Human Thrombopoietin (Thermofisher, cat#PHC9513) and 100ng/mL Recombinant Human Flt-3 Ligand (Thermofisher, cat#CTP9413). Then, we differentiated cells into erythroid cells using a three-step differentiation protocol (Giarratana, et al, 2005). In brief, we cultured CD34+ cells for 7 days in Step 1 media, consisting of Iscove’s modified Dulbecco’s medium (IMDM) (Invitrogen, cat#31980-030) supplemented with 1X GlutaMAX, 100U/mL penicillin-streptomycin, 5% human AB+ plasma (Stemcell Technologies, cat#70039.6), 330ug/mL human holo-transferrin (Sigma, cat#T0665), 10ug/mL human insulin (Sigma, cat#I9278), 2U/mL heparin (Sigma, cat#H3149), 1uM/mL hydrocortisone (Sigma, cat#H6909), 3U/mL recombinant human erythropoietin (EPO) (Invitrogen, cat#PHC2054), 100ng/mL SCF and 5ng/mL interleukin 3 (IL-3) (Sigma, cat#H7166). On day 7, we transferred cells to Step 2 media, a Step 1 media without hydrocortisone and IL-3, and cultured them for 3 to 4 days. We then transferred cells to a Step 3, which has the same composition as Step 2 media without SCF. We cultured cells in Step 3 media for 8 to 9 days. We purchased CD34+ cells from healthy donors from Stemcell Technologies (cat#70002). We isolated CD34+ cells from sickle cell blood donors by CD34+ cell positive selection on PBMC using magnetic beads coupled with an antibody anti-CD34 from Miltenyi Biotec (cat#130-100-453). PBMC were previously isolated by Ficoll gradients from whole blood.

### HUDEP-2 cell culture and differentiation

HUDEP-2 cells were cultured in maintenance media consisting of StemSpan Serum-Free Expansion Medium (SFEM) (Stemcell Technologies, cat#09600), supplemented with 50ng/mL SCF (Thermofisher, cat#CTP2113), 1µM dexamethasone (Millipore, cat#265005,), 1µg/mL doxycycline (Clonetech, cat#631311), 3IU/mL EPO (Invitrogen, cat#PHC2054) and 1% penicillin-streptomycin. For differentiation, cells were cultured in IMDM media supplemented with 1mg/mL of holo-transferrin (Sigma, cat#T0665), 10ug/mL Insulin (Sigma, cat#I9278), 3U/mL heparin (Sigma, cat#H3149), 2% FBS (FBS (Fisher Scientific, cat#SH3007003)), 3% Human Serum Albumin (Irvine Scientific, cat#9988), 1µg/mL doxycycline, 50ng/mL SCF, 3IU/mL EPO and 1% penicillin-streptomycin.

### Flow cytometry

To determine the percentage of HbF-positive cells (F-cells), we fixed cells with glutaraldehyde (Sigma, cat#G5882) and permeabilized differentiated cells with Triton (Life Technologies, cat#HFH10). We stained cells with phycoerythrin (PE)Lconjugated anti-CD235 antibody (ThermoFisher, cat#12-9987-82) and PE-Cy7-conjugated anti-CD71 (Thermofisher, cat#11-0719-42). We measured HbF level using allophycocyanin (APC)-conjugated anti-HbF antibodies (ThermoFisher, cat#MHFH05). The acquisition of stained cells was performed on BD FACSCanto™ and the analysis was run using FlowJo™ Software. To determine the enucleation rate of the erythroid differentiated cells at day 21, we stained cells using living cells marker NucRed (Thermofisher, cat#R37113).

### Western blots

We generated total cell lysates and determined total protein concentration using a BCA protein assay kit (ThermoFisher, cat#23225). Reduced and denatured protein (40μg) was loaded and separated by SDS-PAGE (12% gel), blotted on nitrocellulose membranes (BioRad, cat# 1704159) and finally incubated with antibodies anti-MRP1 (Thermofisher, cat#PIMA516079), anti-human g-globins (g^A^Lglobin and g^G^Lglobin) (Cell Signaling, cat#39386S), anti-Band-3 (Cell Signaling, cat#23276S), anti-LRF (Cell Signaling, cat#50565S), anti-ALAS2 (Abcam, cat#ab244239), anti-GATA-1 (Cell Signaling, cat#3535S), anti-α-tubulin (Cell Signaling, cat#2125S) and anti-β-Actin (Cell Signaling, cat#3700S). Proteins α-tubulin or β-actin served as an internal control. We visualized immunoreactive proteins by using an ECL® (enhanced chemiluminescence) detection system (BioRad, cat#1705062). We measured optical density using ImageJ software (National Institutes of Health, Bethesda, MD)

### Sickling assay

We measured sickling of day 21 fully differentiated erythroid cells from peripheral blood mononuclear cell (PBMC) CD34+ cells from SCD blood donors under hypoxia using a sickle cell imaging flow cytometry assay (SIFCA). Whole blood was collected from adult (>18 years) SCD donors at steady state who had not received a blood transfusion within 3 months following informed consent prior to the blood draw under a Boston Medical Center IRB approved protocol. We challenged SCD donor CD34+ cells under hypoxic conditions by suspending 5×10^5^ cells in 400μL of Hemox buffer (TCS Sci Corporation, cat#HS-500) and incubating under 2% oxygen for 4 hours in a 24-well plate with a control plate incubated under normoxia. After incubation, we fixed cells with 40μL of 25% EM grade freshly thawed glutaraldehyde (Sigma, cat#G5882) and maintained them in their original conditions of hypoxia or normoxia for an additional 20 min. Cells were washed with PBS before analyzing. We sorted and acquired cell images using an Amnis ImageStream X (Luminex) and we quantified shape change using IDEAS software (Luminex). Additional images were obtained using a Zeiss microscope at magnification 20x.

### HPLC

To quantify hemoglobins by high performance liquid chromatography (HPLC), we lysed human CD34+ cells in MilliQ H_2_O and centrifuged hemolysates. We measured hemoglobin variants HbF and HbA_0_ by cation exchange HPLC on a Waters Acquity system with a Thermo ProPac WCX-10 Analytical Column (4 × 250 mm). We eluted proteins in 20mM bisLtris pH 6.95 using a 0 to 200mM NaCl gradient and integrated hemoglobin peak absorbance areas at 410nm.

### *ABCC1* knockout cell generation

We performed CRISPR-mediated knockout of *ABCC1* in HUDEP-2 and CD34+ cells. For both CD34+ and HUDEP-2 cells, we transfected Cas9-guide RNA (gRNA) complexes at an early stage of differentiation (day 2). A synthetic Cas9 was purchased from Aldevron (cat#9214). We acquired *ABCC1* gRNAs from Synthego Corporation with the following sequences: gRNA-1 – TATCTCTCCCGACATCACCG– / gRNA–2 –TTCAGAACACGGTCCTCGTG / gRNA-4 – CGACGTGTCCTCCTTGTTTA–/ gRNA-6 –CCTTGGAACTCTCTTTCGGC–. As a positive control for HbF induction, the following *BCL11A* gRNA was used (Synthego): – TGAACCAGACCACGGCCCGT. To transfect the cells with the complex Cas9-gRNA, we used an electroporation kit optimized for primary erythroid cell transfection (Lonza, cat#NC0301987). We confirmed protein knockdown by western blot.

### *ABCC1*-433Ser clone generation

We obtained the *ABCC1*-433Ser homozygote HUDEP-2 clone (“C6”) from Abcam (Fermont, CA, USA). To generate clones homozygous for the 433Ser mutation, HUDEP-2 cells were cultured and electroporated with Cas9 and gRNA -CGCTCAGAGGTTCATGGACT-, along with a single-stranded oligodeoxynucleotides (ssODN) with sequence AAGGATGACTTGCAGGGGGGCTGACCAGATCATGTTAATGTACGTGGCaAgGTCCATaAAgCTCTGAGCGTCCACAGACATGAGGTTGACAATCTCCCCGACCGTGGAGGA 3’. Editing and homology directed repair (HDR) was confirmed by next generation sequencing (NGS). Single cell clones were derived and screened using NGS. We identified clones C6, D2 and E5, homozygous for the Arg433Ser (AGG>AGC) mutation.

### Giemsa staining

We conducted a Giemsa staining to assess the cell morphology at day 14 of differentiation using a Giemsa Stain kit (May-Grunwald) from Scytek laboratories (cat#GMG-2-IFU). The staining was performed according to the manufacturer protocol.

### Glutathione assay

To measure intracellular glutathione, we used a luminescent-based assay to detect and quantify total glutathione (GSH+GSSG) according to the manufacturer protocol (Promega, cat#V6611).

### RNAseq analysis

We cultured and differentiated for 7 days edited and wild-type HUDEP-2 from 3 independent differentiation experiments, and DMSO or MK571-treated CD34+ cells from 3 healthy donors for 14 days. When the differentiation was completed, we extracted RNA using a Trizol reagent kit (Invitrogen).

We assessed RNA quality on an Agilent TapeStation and quantified RNA by Qubit RNA assay. Poly-A selection and paired-end 2×150bp library preparation (Illumina) was performed by Genewiz, NJ. Libraries were sequenced on Illumina HiSeq 2500 platforms. We trimmed adapter sequences and nucleotides with poor quality using Trimmomatic v.0.36 (Bolger et al, 2014). We mapped trimmed reads to the GRCh38 human reference genome using STAR v.2.5.2b (Veeneman et al, 2016). We estimated gene level read counts using the R Subread package v.1.5.2, including only unique reads falling within exons.

We used DESeq2 (Love et al, 2014) to identify differentially expressed genes between DMSO and MK571-treated cells or between clone C6 and wild-type HUDEP-2 cells. Differential gene expression analyses were adjusted for different experiment batches for HUDEP-2 cells and for donor sources for CD34+ cells. Genes with a false discovery rate (FDR) adjusted *P*-value <0.05 were considered differentially expressed. We used the Qiagen Ingenuity Pathway Analysis (IPA) software for pathway and upstream regulator analysis, including genes with an adjusted *P*-value < 0.05 and absolute fold change > 1.5.

### Proteomics and phospho-proteomics analysis

We extracted proteins from the same CD34+ cells used for the transcriptomic analysis. Samples preparation, protein digestion, peptides tandem mass tag (TMT) labeling and proteomic analysis were performed by IQ Proteomics (Cambridge, MA, USA). Bioinformatic quantification was performed by IQ Proteomics.

For the proteomics analysis, spectra were searched against the Uniprot Human protein sequence database (2021) with forward and reverse sequences and including common contaminants. Peptide assignments were filtered using a target-decoy approach via linear discriminant analysis. Peptides and proteins were filtered to a 1% FDR. A requirement of signal to noise (SN) ratio > 120 was further applied to peptides. Peptides were collapsed to protein groups via the rule of parsimony. The phospho-proteomics analysis was performed as the proteomics analysis, but including phosphorylation as a variable modification in the search against the Uniprot protein database. In addition to requiring a SN >120, an additional requirement of Isolation Purity > 50% was applied to phosphopeptides. Phosphopeptides were reduced to protein sites based on localization according to a binomial probability.

We performed protein differential expression analysis between DMSO and MK571-treated cells using the R package DEqMS v1.4.0, adjusting for sample donor ID. We identified differentially expressed proteins based on Spectra count adjusted (SCA) *P*-values. We used the Qiagen IPA software for pathway and upstream regulator analysis. For the proteomics pathway analysis, we included proteins with a nominal *P*-value <0.05 and absolute fold change >1.5. For the phospho-proteomics analyses, we included protein with phosphorylation absolute log fold change >1.5 at at least one site. We performed a comparison analysis between the RNAseq, proteomics, and phosphoproteomics experiment using IPA to identify shared pathways modulated by pharmacological inhibition of MRP1. Association between gene expression and protein expression was estimated by Pearson’s correlation in R.

## Data Availability

Upon publication, the INTERVAL HbF GWAS summary statistics will be made publicly available in the NHGRI-EBI GWAS Catalog (https://www.ebi.ac.uk/gwas/) and the RNA sequencing data will be available via the Gene Expression Omnibus portal (https://www.ncbi.nlm.nih.gov/geo/).

## ACKNOWLEDGMENTS

The fetal hemoglobin measurements in INTERVAL were funded by Biogen. Experimental follow-up was supported by a grant from the Sanofi *iAwards* Europe Program.

Participants in the INTERVAL randomised controlled trial were recruited with the active collaboration of NHS Blood and Transplant England (www.nhsbt.nhs.uk), which has supported field work and other elements of the trial. DNA extraction and genotyping were co-funded by the National Institute for Health and Care Research (NIHR), the NIHR BioResource (http://bioresource.nihr.ac.uk) and the NIHR Cambridge Biomedical Research Centre (BRC-1215-20014) [*]. The academic coordinating centre for INTERVAL was supported by core funding from the: NIHR Blood and Transplant Research Unit in Donor Health and Genomics (NIHR BTRU-2014-10024), NIHR BTRU in Donor Health and Behaviour (NIHR203337), UK Medical Research Council (MR/L003120/1), British Heart Foundation (SP/09/002; RG/13/13/30194; RG/18/13/33946) and NIHR Cambridge BRC (BRC-1215-20014) [*]. A complete list of the investigators and contributors to the INTERVAL trial is provided in reference [**]. The academic coordinating centre would like to thank blood donor centre staff and blood donors for participating in the INTERVAL trial.

Professor John Danesh holds a British Heart Foundation Professorship and a NIHR Senior Investigator Award [*].

This work was supported by Health Data Research UK, which is funded by the UK Medical Research Council, Engineering and Physical Sciences Research Council, Economic and Social Research Council, Department of Health and Social Care (England), Chief Scientist Office of the Scottish Government Health and Social Care Directorates, Health and Social Care Research and Development Division (Welsh Government), Public Health Agency (Northern Ireland), British Heart Foundation and Wellcome.

The authors would like to thank RIKEN institute (Japan) for providing the HUDEP-2 cells, William Kuhlman from Sanofi for helping with HPLC analysis, and colleagues at the University of Cambridge and the National Haemoglobinopathy Reference Laboratory for their help with preparing and shipping INTERVAL samples and conducting HbF assays.

*The views expressed are those of the author(s) and not necessarily those of the NIHR, NHSBT or the Department of Health and Social Care.

**Di Angelantonio E, Thompson SG, Kaptoge SK, Moore C, Walker M, Armitage J, Ouwehand WH, Roberts DJ, Danesh J, INTERVAL Trial Group. Efficiency and safety of varying the frequency of whole blood donation (INTERVAL): a randomised trial of 45 000 donors. Lancet. 2017 Nov 25;390(10110):2360-2371.

