## Supplementary Figures for "Genomic discovery and functional validation of MRP1 as a novel fetal hemoglobin modulator and potential therapeutic target in sickle cell disease"

**Supplementary Tables**

Supplementary Table 1: Characteristics of INTERVAL participants

Supplementary Table 2: Sentinel variants at ten significant loci from the unconditioned GWAS.

Supplementary Table 3: Associations of HbF variants with other blood cell traits.

Supplementary Table 4: List of 30 red blood cell traits examined in INTERVAL.

Supplementary Table 5: Mediation analysis of HbF associations with blood cell traits.

Supplementary Table 6: Association of *ABCC1* 433S with plasma metabolites in the INTERVAL study.

Supplementary Table 7: Phenome-wide association scan for *ABCC1* 433S.

Supplementary Table 8: Genes differentially expressed between erythroid-differentiated HUDEP-2 *ABCC1* 433S clone C6 and wild-type cells.

Supplementary Table 9: Enrichment analysis for differentially expressed genes in HUDEP-2 *ABCC1* 433S clones compared to wild-type.

Supplementary Table 10: Genes differentially expressed between erythroid-differentiated human CD34+ cells treated with MK571 or DMSO.

**Supplementary Figures**

Supplementary Figure 1: Quantile-quantile plot for the unconditioned GWAS of HbF levels.

Supplementary Figure 2: Manhattan plot of the unconditioned GWAS indicating ten regions with genome-wide significant association signals.

Supplementary Figure 3: Regional association plots for the *ABCC1* region with HbF levels, plasma protein and metabolite levels in INTERVAL.

Supplementary Figure 4: Phenome-wide association scan for *ABCC1* p.R433S (rs60782127).

Supplementary Figure 5: CRISPR-Cas9 mediated *ABCC1* 433Ser knock-in leads to increased fetal hemoglobin in HUDEP-2 cells.

Supplementary Figure 6: CRISPR-Cas9 mediated *ABCC1* knockout leads to increased fetal hemoglobin in HUDEP-2 cells.

Supplementary Figure 7: CRISPR-Cas9 mediated *ABCC1* knock-out leads to increased HbF production in erythroid cells derived from primary CD34+ HSCs.

Supplementary Figure 8: Inhibition of MRP1 by MK571 does not impair erythroid differentiation of CD34+ primary HSCs.

**Supplementary Figures**

Supplementary Figure 1: Quantile-quantile plot for the unconditioned GWAS of HbF levels.

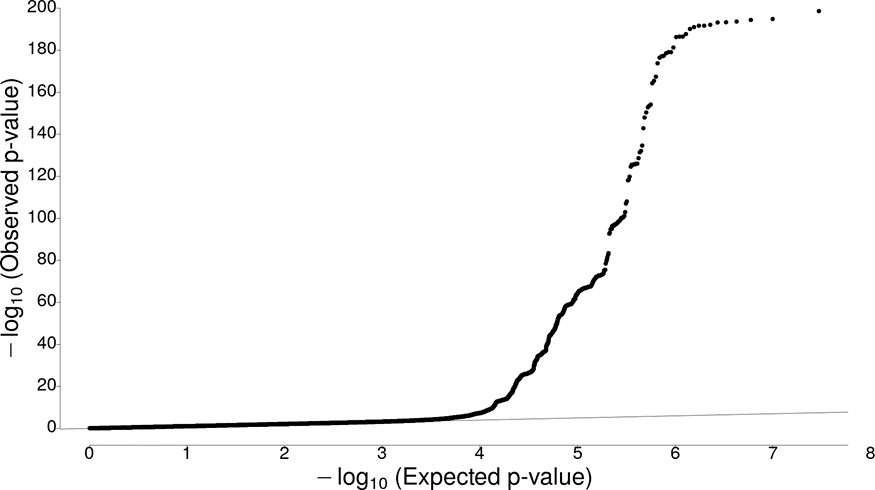

*Estimated genomic control inflation factor was 1.047.*

Supplementary Figure 2: Manhattan plot of the unconditioned GWAS indicating ten regions with genome-wide significant association signals.

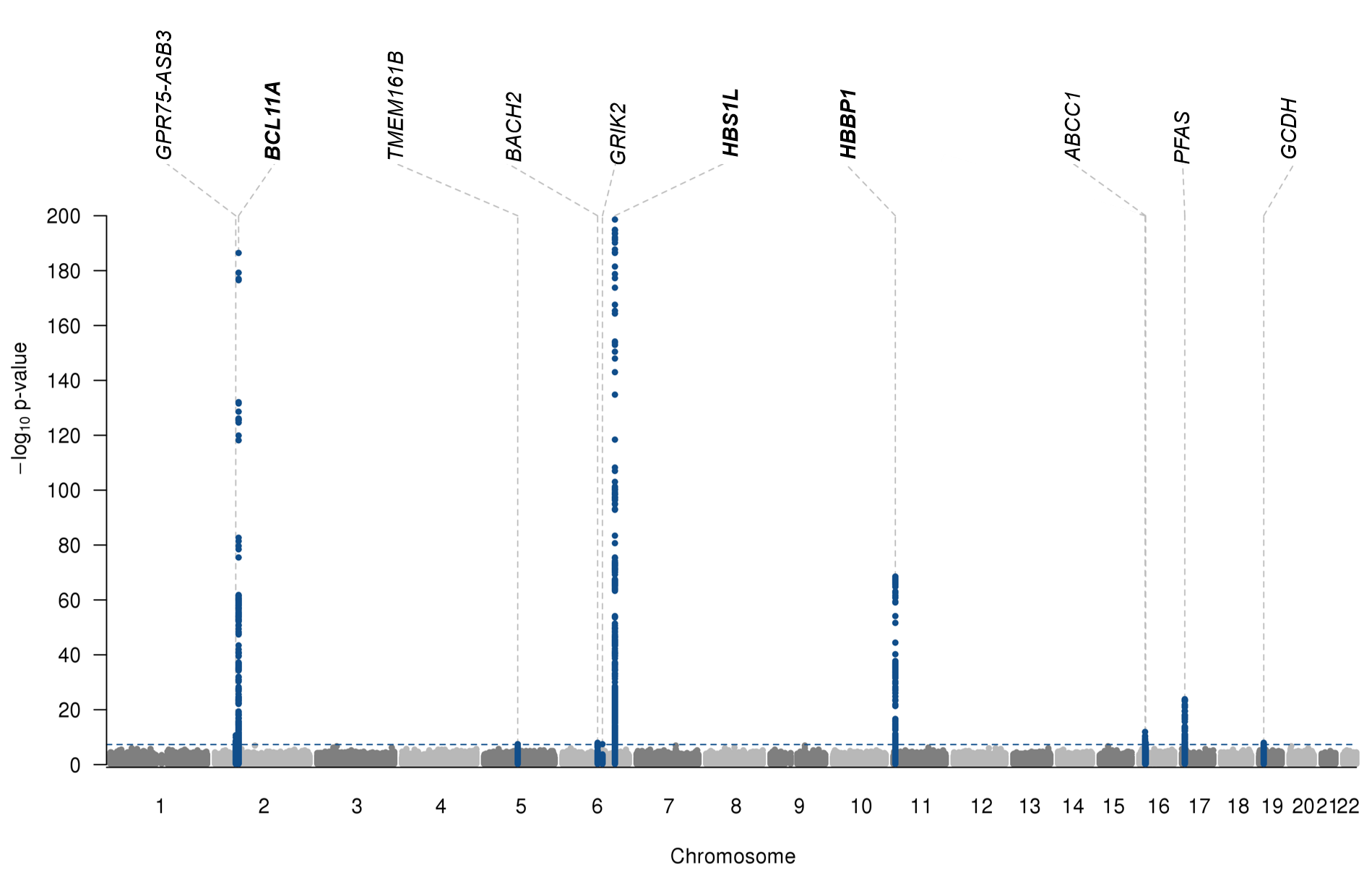

*Loci reaching genome-wide significance in the unconditioned GWAS are denoted in blue. Gene names denote nearest genes to the lead variant at each locus. Gene names in bold font denote previously reported loci. (The ARGHAP39 locus on chromosome 8 was only significant after conditional analysis).*

Supplementary Figure 3: Regional association plots for the *ABCC1* region with HbF levels, plasma protein and metabolite levels in INTERVAL.

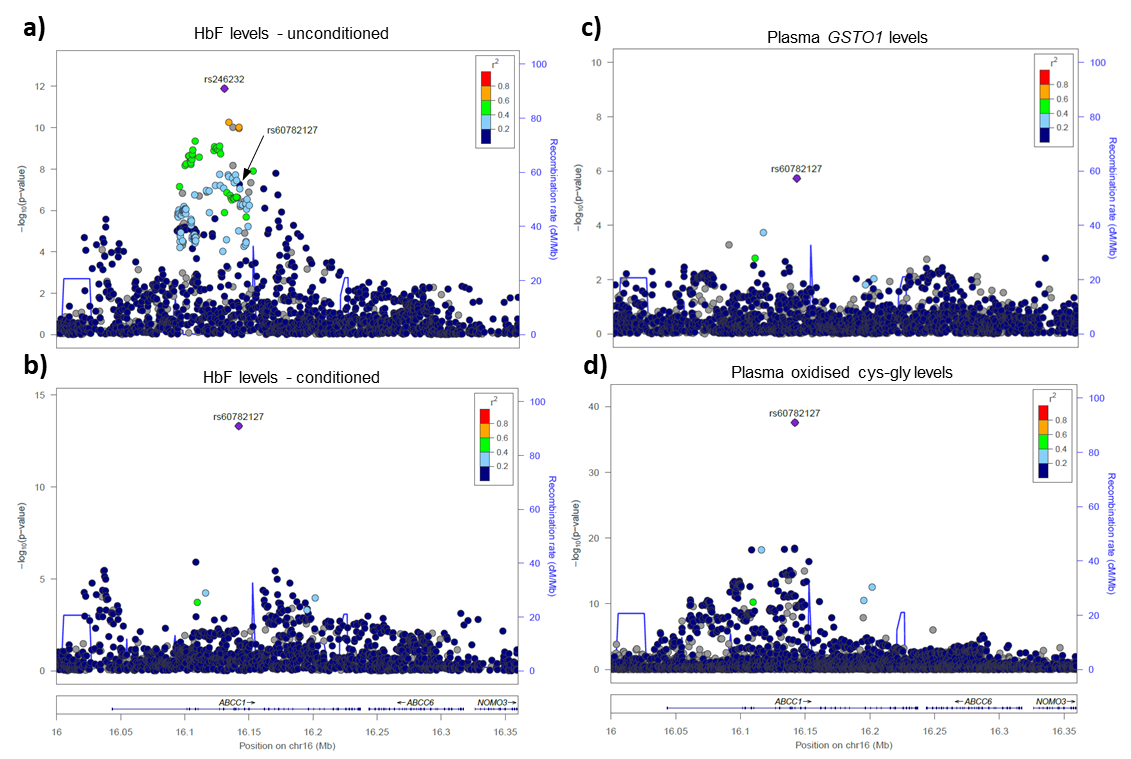

*a) Regional association plot for the ABCC1 region with circulating HbF levels in the INTERVAL study. The two sentinel variants (rs246232 and rs60782127) for the two independent signals are labelled.*

*b) Regional association plot for the ABCC1 region with circulating HbF levels in the INTERVAL study after conditioning on the lead variant rs246232. The independent signal with rs60782127 (p.Arg433Ser) is labelled.*

*c) Regional association plot for the ABCC1 region with plasma GSTO1 levels in the INTERVAL study, measured using the SomaLogic SomaScan aptamer-based assay.*

*d) Regional association plot for the ABCC1 region with plasma oxidised cys-gly levels in the INTERVAL study, measured using the Metabolon HD4 mass-spectrometry assay. Plots for other related metabolites looked similar but are not shown.*

Supplementary Figure 4: Phenome-wide association scan for *ABCC1* p.R433S (rs60782127).

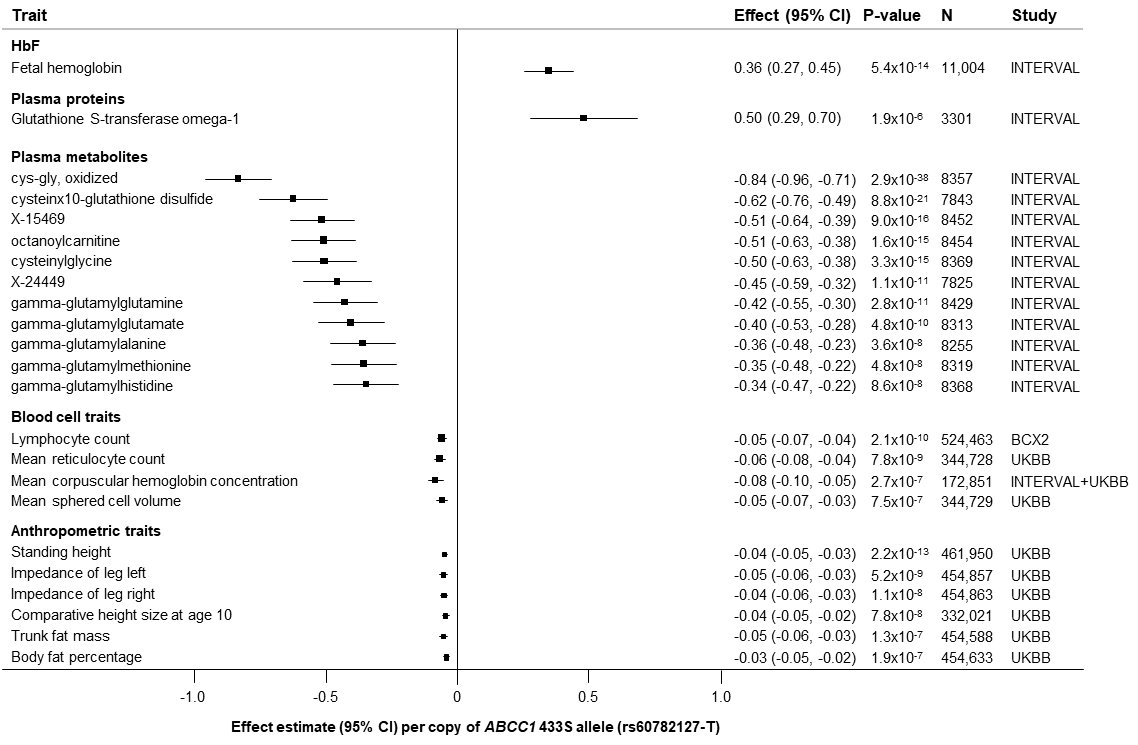

Supplementary Figure 5: CRISPR-Cas9 mediated *ABCC1* 433Ser knock-in leads to increased fetal hemoglobin in HUDEP-2 cells.

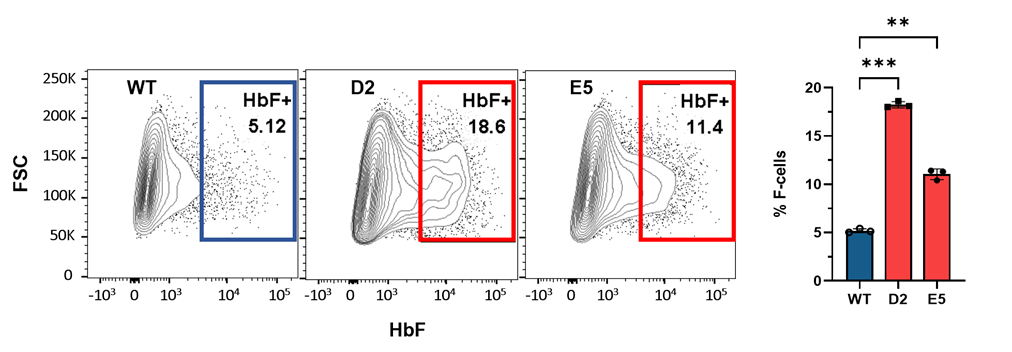

*Homozygous knock-in of the ABCC1 433Ser allele in HUDEP-2 clones D2 (N=3) and E5 (N=3) leads to increased HbF levels compared to wild-type cells (N=3) as quantified by flow cytometry after 7 days of erythroid differentiation (right). Representative flow cytometry plots from a single experiment (left). FSC: Forward scatter*. *Two-tailed t-test, **P<0.01, ***P<0.001 (N=3).*

Supplementary Figure 6: CRISPR-Cas9 mediated *ABCC1* knockout leads to increased fetal hemoglobin in HUDEP-2 cells.

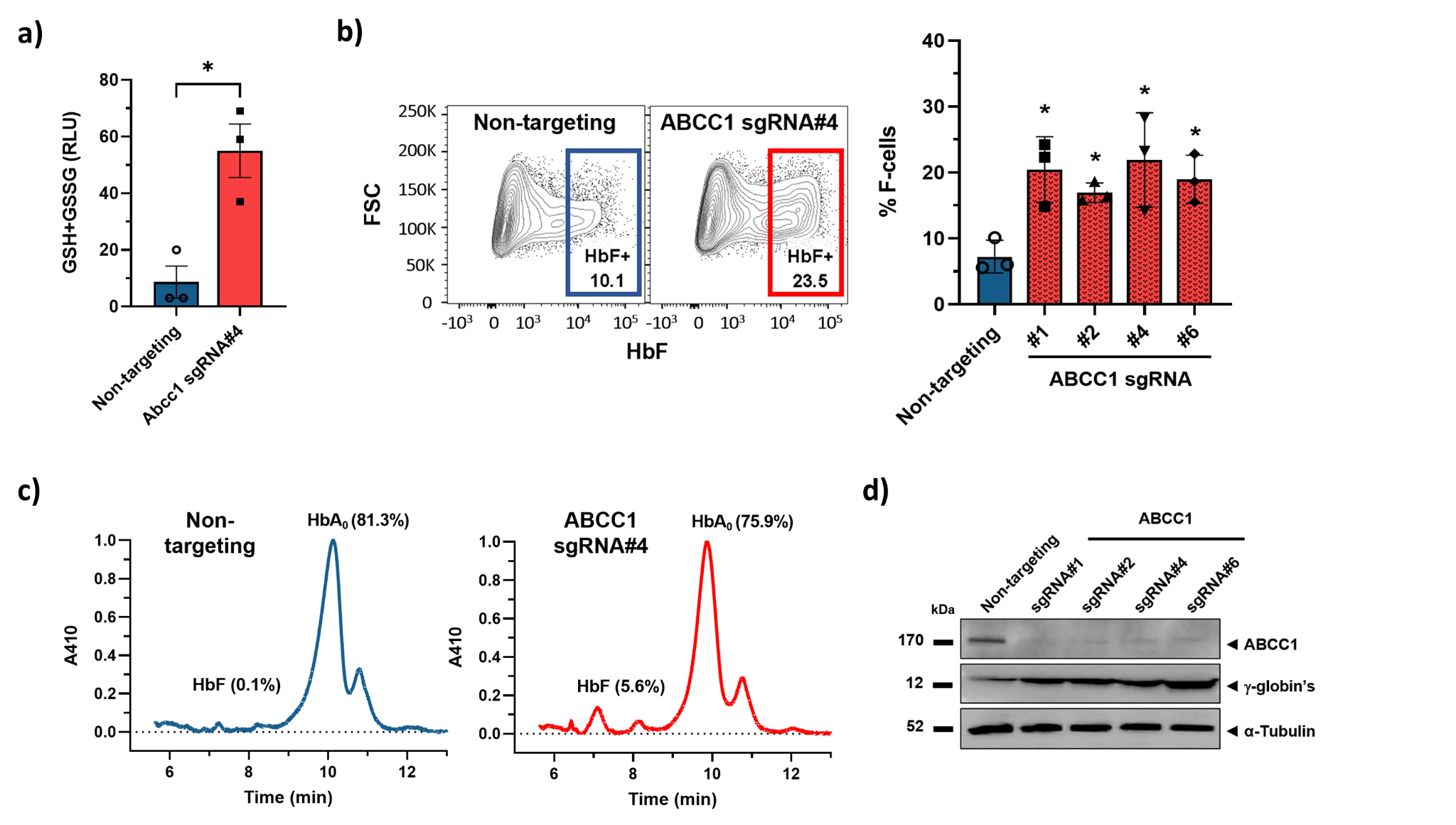

*a) Measurement of total intracellular glutathione (GSH+GSSG) in ABCC1-KO HUDEP-2 cells shows an increase compared to control. Two-tailed t-test, *P<0.05 (N=3).*

*b) ABCC1 CRISPR-Cas-9 mediated knockout with sgRNA#1, #2, #4 and #6 significantly increases HbF‑positive cells at 7 days of differentiation in HUDEP-2 cells compared to control (non-targeting) quantified by flow cytometry (right). Panels are representative flow cytometry plots (left) from a single donor.*

*c) Fetal hemoglobin induction in ABCC1-KO HUDEP-2 cells was confirmed by cation-exchange high-performance liquid chromatography (HPLC).*

*d) Representative western blot of differentiating ABCC1-KO HUDEP-2 cells confirms ABCC1 silencing and gamma-globin induction. Similar results were obtained for three independent donors.*

Supplementary Figure 7: CRISPR-Cas9 mediated *ABCC1* knock-out leads to increased HbF production in erythroid cells derived from primary CD34+ HSCs.

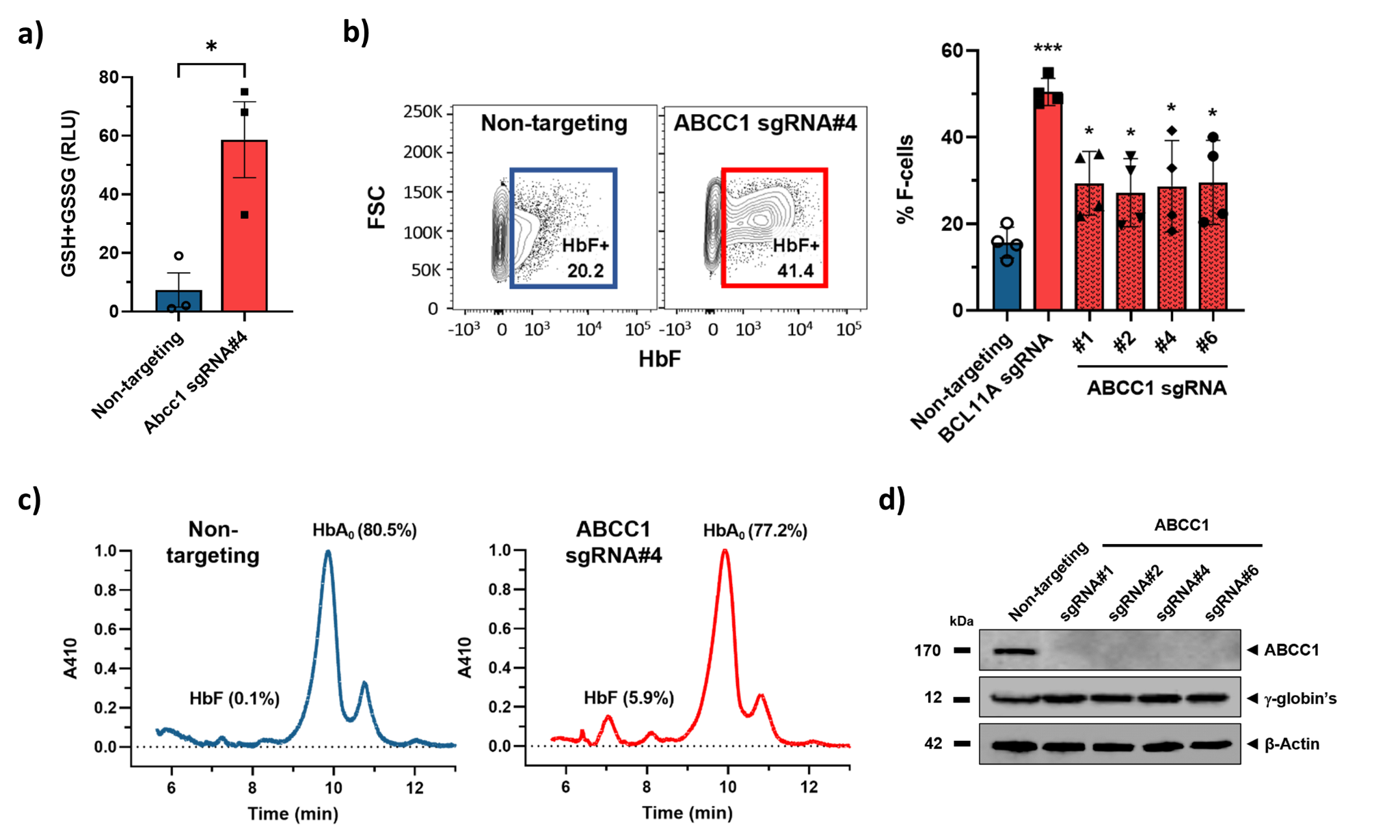

*a)* *Measurement of total intracellular glutathione (GSH+GSSG) in ABCC1-KO CD34+ cells shows an increase compared to control and confirms ABCC1 silencing. Two-tailed t-test, *P<0.05, **P<0.01 (N=3).*

*b) ABCC1 silencing with sgRNA#1, #2, #4 and #6 by CRISPR-Cas-9 gene editing significantly increases HbF‑positive cells by day 14 of differentiation in CD34+ cells from healthy donors compared to control (non-targeting) quantified by flow cytometry (right). Panels are representative plots from a single donor (left).*

*c) Fetal hemoglobin induction in ABCC1-KO CD34+ cells was confirmed by cation-exchange high-performance liquid chromatography (HPLC).*

*d) Representative western blot of differentiating CD34+ cells at day 14 confirms ABCC1 silencing and fetal hemoglobin induction. Similar results obtained for three independent donors.*

Supplementary Figure 8: Inhibition of MRP1 by MK571 does not impair erythroid differentiation of CD34+ primary HSCs.

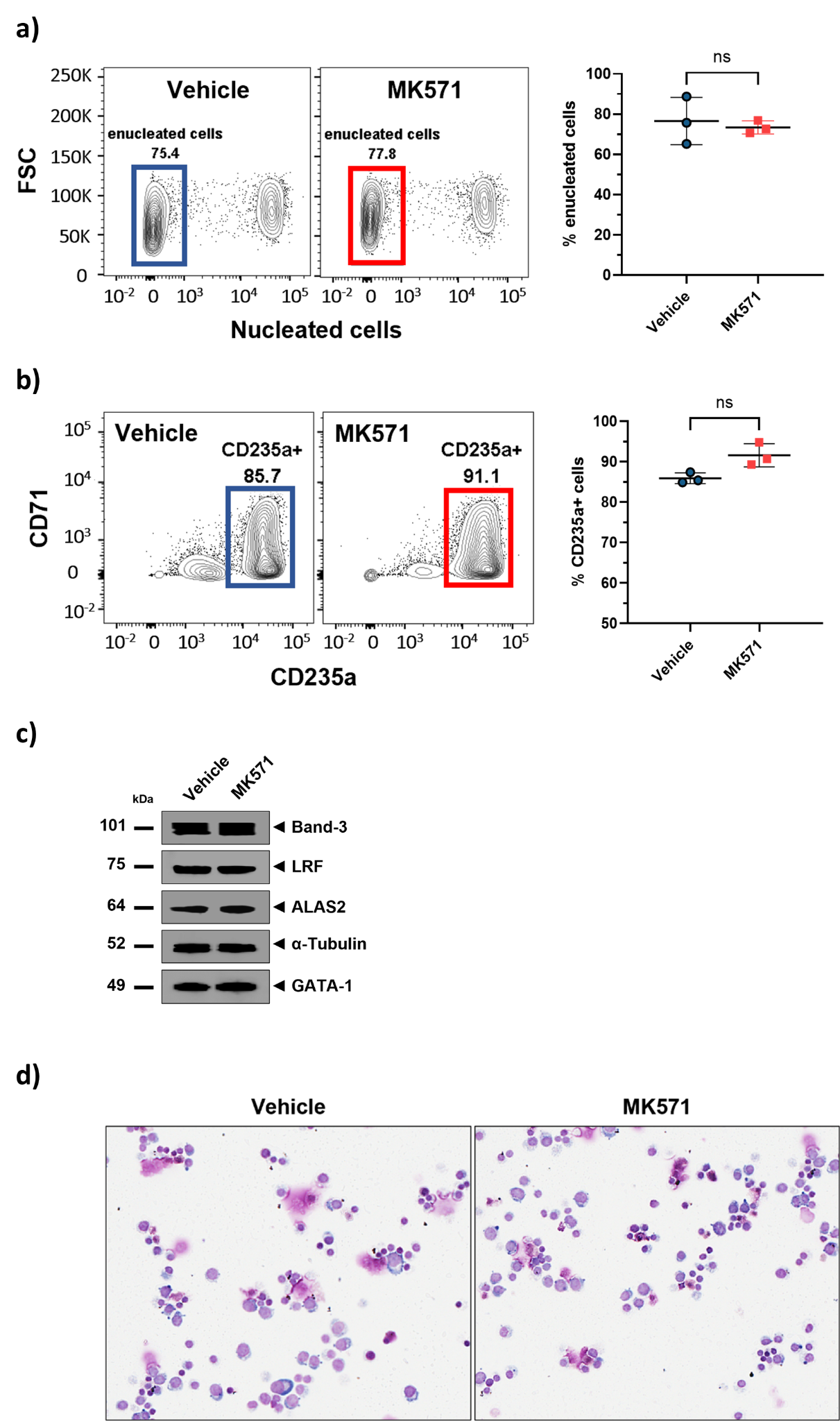

*a) High levels of enucleation observed for fully differentiated CD34+ cells on day 21 for both MK571 and vehicle treatments confirms normal erythroid differentiation. Panels are representative flow cytometry plots from a single donor (right). Two-tailed t-test, NS = not significant (N=3).*

*b) Flow cytometric analysis of CD235a+ cells on day 21 indicates normal differentiation of CD34+ cells from healthy donors when treated with MK571 (50µM). Panels are representative flow cytometry plots from a single donor (left). Two-tailed t-test, NS = not significant (N=3).*

*c) Western blot analysis of additional erythroid differentiation markers (Band-3, LRF, ALAS2, GATA-1) on day 14 shows similar protein expression levels between MK571- and vehicle- treated CD34+ cells after normalization to alpha-tubulin, confirming normal cell erythroid differentiation when MRP1 is inhibited.*

*d) Giemsa staining of differentiated CD34+ cells from healthy donors treated with MK571 (50µM) on day 14 confirms normal cell morphologies following treatment (all magnifications 40X).*
